# Immune disturbances cluster around specific depressive phenotypes under conditions of structural adversity

**DOI:** 10.64898/2026.07.15.26358135

**Authors:** Caio Hofmann Francisco Alves, Fausto Coutinho-Lourenço, Yuan-Pang Wang, Danielle Bivanco-Lima, Isabella Bensenor, Geilson Lima Santana, Bruno Mendonça Coelho, Maria C. Viana, João Maurício Castaldelli-Maia, Livia A. Carvalho, Laura Helena Andrade

## Abstract

**Background:** Depression is clinically heterogeneous, and immune-metabolic disturbances may not map uniformly onto categorical diagnosis or symptom severity. In populations exposed to substantial structural adversity, it remains unclear whether inflammatory and metabolic biomarker differences reflect adversity exposure itself or cluster around specific depressive phenotypes.

**Methods:** Data were drawn from the São Paulo Megacity Mental Health Survey, a population-based study in which 5,037 household residents underwent structured psychiatric interviews. Among 770 participants assessed as having clinically significant symptoms (SCID-I), individuals with chronic physical illnesses, hs-CRP >20 mg/L, or missing data were excluded, yielding an analytic sample of 653. Latent class analysis of 16 DSM-IV depressive symptoms was used to identify symptom-derived phenotypes. Multinomial logistic regression tested associations between latent classes and immune-metabolic biomarkers, including hs-CRP, lipid fractions, fasting glucose, and triglycerides, adjusting for age, sex, education, smoking, and BMI.

**Results:** A four-class solution identified asymptomatic (44.56%), mild-moderate (19.14%), atypical-like (16.69%), and melancholic-like (19.60%) classes. The two high-severity classes diverged by neurovegetative features: atypical-like depression was characterised by weight gain, hypersomnia, and psychomotor retardation, whereas melancholic-like depression was characterised by weight loss, insomnia, and psychomotor agitation. hs-CRP was highest in the atypical-like class and lowest in the melancholic-like class despite similar symptom severity. After BMI adjustment, elevated hs-CRP in the atypical-like class attenuated, whereas lower hs- CRP in the melancholic-like class persisted. Other metabolic markers did not robustly differentiate classes after adjustment.

**Conclusions:** Immune-metabolic differences in depression do not simply follow symptom severity. In this São Paulo cohort, higher and lower hs-CRP profiles clustered around distinct latent depressive phenotypes, supporting biologically heterogeneous depressive presentations within a socially exposed urban population.

**Highlights:**

- Latent class analysis (LCA) of 16 DSM-IV symptoms identified four symptom-derived classes: asymptomatic, mild–moderate, atypical-like, and melancholic-like.
- The two high-severity classes diverged primarily by neurovegetative features: weight gain/hypersomnia versus weight loss/insomnia.
- hs-CRP was highest in the atypical-like class and lowest in the melancholic-like class.
- Elevated hs-CRP in the atypical-like class attenuated after BMI adjustment, whereas lower hs-CRP in the melancholic-like class persisted after BMI adjustment.
- Findings suggest that immune-metabolic differences in depression may cluster around symptom-derived phenotypes rather than severity alone.

## Introduction

Major depressive disorder (MDD) is one of the leading contributors to disability worldwide (1). Despite advances in pharmacological and psychological treatments, a substantial proportion of patients fail to respond adequately, underscoring marked heterogeneity in symptom presentation, course, and biology (2). The current diagnostic definition of depression, in which any five of nine symptoms may be sufficient for diagnosis, permits numerous symptom constellations, such that two patients can share the same diagnosis with minimal symptom overlap (3). This raises the possibility that MDD aggregates biologically and clinically distinct phenotypes rather than representing a single pathophysiological entity.

This question is particularly relevant in São Paulo, one of the world’s largest megacities and a setting characterised by substantial structural and environmental adversity. The burden of MDD is elevated in Latin America, where recent regional analyses estimate a prevalence of approximately 12%, more than twice the global average (1). In São Paulo, urban stressors including violence exposure, social deprivation, and limited green-space availability are key contextual features that may contribute to both depression risk and depression heterogeneity. In this context, structural adversity is not merely a background exposure, but a setting in which biological differentiation between depressive phenotypes can be tested. Understanding whether immune–metabolic disturbances cluster around specific symptom profiles in such a population may help clarify whether biomarker associations reflect depression severity, adversity-linked distress, or more specific clinical phenotypes.

Immune and metabolic disturbances have been implicated in the aetiology and pathophysiology of depression (15,16). Altered levels of pro-inflammatory cytokines and enhanced expression of immune-related genes have been reported in the periphery and brain of patients with severe mood disorders (17,18). However, immune disturbances are not uniformly elevated across all individuals with depression, and associations between immune-metabolic biomarkers and categorical psychiatric diagnoses are often heterogeneous (19–25). Immune-metabolic disturbances may predict the development of depressive symptoms in previously healthy individuals, and Mendelian randomisation analyses suggest a probable causal role for interleukin-6, CRP and triglycerides (26). Anti-inflammatory interventions have also been shown to reduce depressive symptoms in some clinical contexts but is not uniformly beneficial (27,28). These findings support immuno-metabolic models of depression, in which chronic psychological stress and related biological pathways influence immune-metabolic dysregulation, and brain disfunction (29,30). However, they also suggest that immune–metabolic dysfunction may characterise only specific depressive phenotypes rather than all individuals with depression.

Data-driven approaches may help parse this heterogeneity by identifying symptom-derived phenotypes that are more clinically and biologically coherent than categorical diagnoses alone (6). Latent class analysis (LCA) is one such approach and has previously been used to identify depressive subtypes, including atypical and non-atypical presentations (7–14). Here, we used LCA of DSM-IV depressive symptoms in the São Paulo Megacity Mental Health Survey to identify latent depressive phenotypes and test whether these classes differ in immune– metabolic biomarkers. We were particularly interested in whether biomarker differences would reflect a simple gradient of depression severity, or whether immune–metabolic profiles, including both higher and lower biomarker levels, would cluster around specific clinical phenotypes within a population exposed to substantial structural adversity (31, 60).

## Methods

### Ethics

Procedures for recruitment, consent, and protection of human subjects in the São Paulo Megacity Mental Health Survey were approved by the Research and Ethics Committee of the University of São Paulo Medical School (Project 792/03). All participants provided written informed consent.

### Sample characteristics and study design

The São Paulo Megacity Mental Health Study (32) is a population-based cross-sectional study of adults (≥18 years) residing in non-institutionalized households in the São Paulo Metropolitan Area, the largest Brazilian metropolis. Data were collected between May 2005 and April 2007 using a stratified, multistage, clustered area probability design. In the Household Phase (n=5,037), participants completed sociodemographic and health assessments and the WMH-CIDI (Brazilian-Portuguese adaptation).

In the Clinical Phase, all respondents with any WMH-CIDI mental disorder and a subsample without disorders were invited for in-person assessment at the Institute of Psychiatry (IPq-HCFMUSP). A total of 770 individuals were evaluated by trained psychiatrists using the SCID-I for DSM-IV. Two physicians conducted physical examinations and obtained medical history, including cardiovascular and cerebrovascular disease and diabetes. For the present analysis, we excluded 70 participants with chronic physical illnesses and 17 with hs-CRP >20 mg/L (indicative of acute inflammation). We restricted to participants with complete SCID depressive symptoms, hs-CRP, and anthropometric data, yielding a final analytic sample of 653.

To evaluate potential selection bias between the Household and Clinical Phase, we compared the analytic subsample with the full household cohort. The subsample was younger, had higher education, and included a higher proportion of women—consistent with enrichment for psychiatric morbidity and clinic attendance (Supplementary Table S1).

### Measures

#### Psychiatric clinical interview

Experienced psychiatrists administered the SCID-I (Brazilian-Portuguese) to ascertain DSM-IV disorders (32,35). The MDD section comprised 27 items assessing current or worst-lifetime episode symptoms. Responses (absent/uncertain/present) were dichotomized; no “uncertain” ratings occurred. Redundant items (e.g., separate queries for weight gain vs weight loss after a general weight-change item) were removed, leaving 19 dichotomous symptoms: sadness, irritable mood, loss of interest, weight gain, weight loss, insomnia, hypersomnia, psychomotor retardation, psychomotor agitation, fatigue, worthlessness, guilt, concentration problems, indecision, thoughts of death, suicidal ideation, suicide plan, suicide attempt, and loss of functioning. Global severity was rated with the Clinical Global Impression–Severity (CGI-S).

#### Blood biomarkers

Morning fasting venous blood was drawn at Instituto de Psiquiatria, Hospital das Clínicas, Faculdade de Medicina, Universidade de São Paulo (IPq-HCFMUSP) using standard phlebotomy after a 12-hour fast. Assays included high-sensitivity C-reactive protein (hs-CRP; nephelometry), glucose (hexokinase), total cholesterol (enzymatic colorimetry), HDL-cholesterol (homogeneous assay), and triglycerides (enzymatic colorimetry). LDL-cholesterol was calculated via the Friedewald formula (32). Analyses were performed on a Konelab 60i platform. Lipid profile, glucose levels, and inflammation are established components of allostatic load indices.

#### Health and sociodemographic characteristics

Covariates were sex (female/male), age (years), education (years), smoking (current vs non-smoker), height (m) and weight (kg) measured per CDC standardized procedures, and body mass index (BMI, kg/m²).

### Statistical analysis

Latent Class Analysis (LCA): We first screened the 19 symptoms for critically low endorsement, discrimination, and communality to improve interpretability and model stability. Suicidal ideation (∼7% endorsement), plan (∼0.5%), and attempt (∼2.5%) showed very low information value, commonality and risk of quasi-complete separation increasing model instability and were thus removed prior to the final LCA, leaving 16 indicators. LCA was conducted in R with the poLCA package (46) using maximum-likelihood estimation. To ensure convergence for the most successful LCA models, we used 500 random starts and 10 final optimizations in the final stage of convergence. Competing models (2–8 classes) were compared using log-likelihood, AIC, BIC, Pearson χ² goodness-of-fit, deviance (G²), and relative entropy. Model selection prioritized parsimony (minimum BIC), adequate cell counts (≤10–20% of cells with <5 observations), and clinical interpretability.

Associations with biomarkers: We fit multinomial logistic regressions with latent class factors as the outcome. CRP and triglycerides were log-transformed due to skew. Model 1 adjusted for age, sex, education, and smoking; Model 2 additionally adjusted for BMI to evaluate adiposity’s role. Descriptive statistics were used for sample characterization. Analyses were conducted in R 4.1.3.

## Results

### Latent class model selection

Identification of depressive subtypes: Fit indices for 2–8 class solutions are summarized in Table 1. Entropy values were modest across models, ranging from 0.48–0.53. Models with three- to five- classes minimized BIC values, the five-class model was not retained because the smallest class contained fewer than 10% of individuals. The 4-class model provided the best overall balance of parsimony (lowest BIC), statistical fit (χ²), class size, and clinical interpretability.

**Table 1.** Latent Class Analysis model indices.

| **Model** | **LL** | **AIC** | **BIC** | **(Deviance)** | **(Chi-square)** | **Relative Entropy** | **df** | **% smallest class** |
| --- | --- | --- | --- | --- | --- | --- | --- | --- |
| Two-class | -3843.65 | 7753.31 | 7901.20 | 2488.31 | 77855.71 | 0.53 | 620 | 47.63% |
| Three-class | -3685.55 | 7471.10 | 7695.18 | 2172.11 | 47551.21 | 0.51 | 603 | 19.45% |
| Four-class | -3610.95 | 7355.91 | 7656.17 | 2022.91 | 40614.38 | 0.50 | 586 | 16.69% |
| Five-class | -3570.64 | 7309.28 | 7685.73 | 1942.29 | 56330.67 | 0.49 | 569 | 6.13% |
| Six-class | -3543.81 | 7289.62 | 7742.26 | 1888.63 | 59139.40 | 0.49 | 552 | 6.58% |
| Seven-class | -3516.57 | 7269.14 | 7797.97 | 1834.15 | 40893.32 | 0.48 | 535 | 5.36% |
| Eight-class | -3494.03 | 7258.07 | 7863.08 | 1789.08 | 30262.01 | 0.48 | 518 | 2.14% |

### Symptom profiles of the four latent classes

The four-class model comprised an asymptomatic class (44.56%), a mild–moderate class (19.14%), and two high-severity classes differentiated by vegetative/psychomotor features: the atypical-like subtype (16.69%) with elevated probabilities of weight gain, hypersomnia, and psychomotor retardation; the melancholic-like class (19.60%) characterized by higher probabilities of weight loss, insomnia, and psychomotor agitation.

Symptom probability profiles for the four-class model are shown in Figure 2. The model separated participants partly by overall symptom burden, but the two higher-severity classes were distinguished by opposing neurovegetative patterns. Most depressive symptoms showed broadly similar probabilities across these higher-severity classes, whereas appetite/weight change, sleep disturbance and psychomotor disturbance showed divergent profiles.

**Figure 1.**
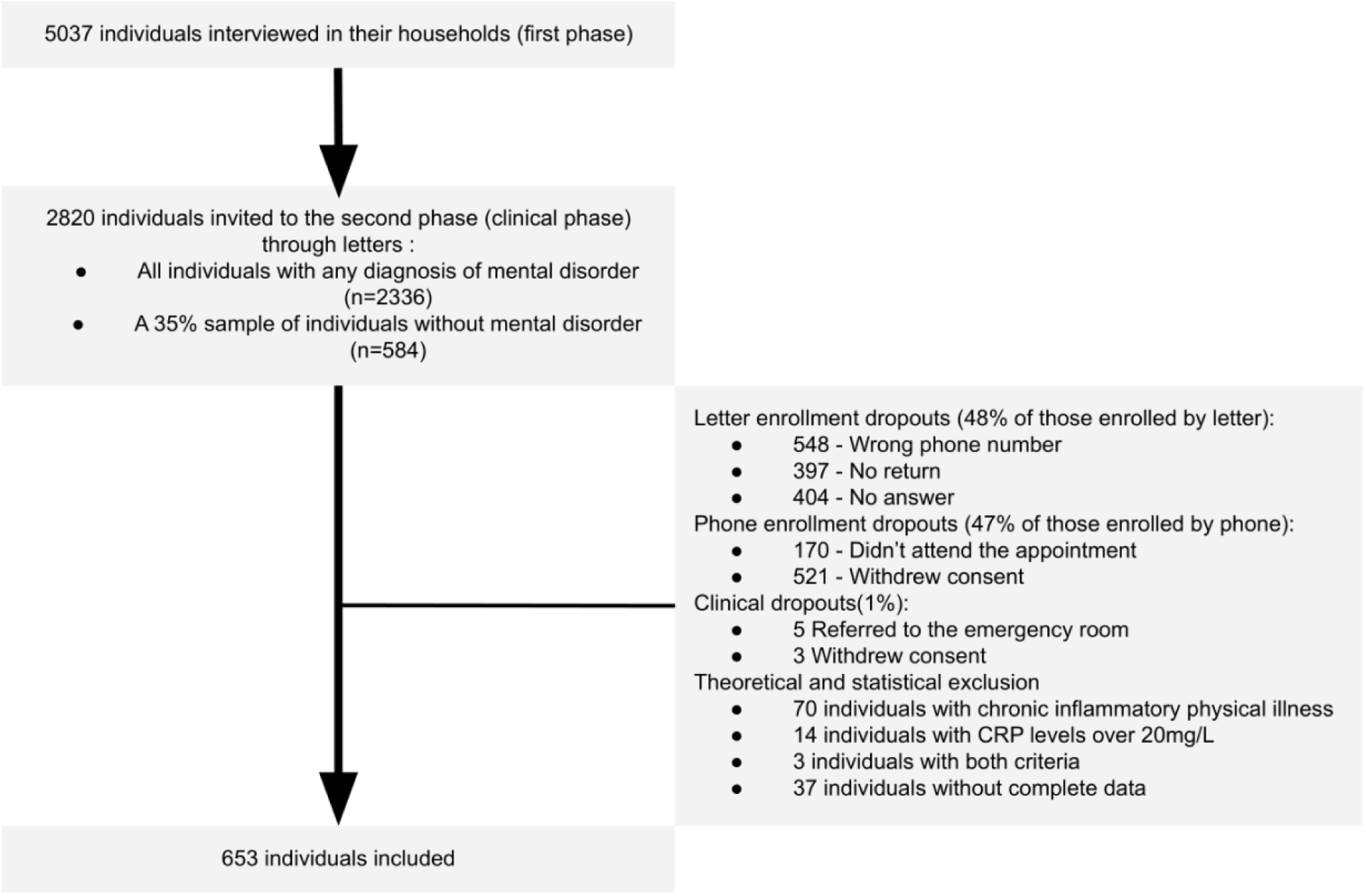
Flowchart of the process employed on data collection and selection.

**Figure 2.**
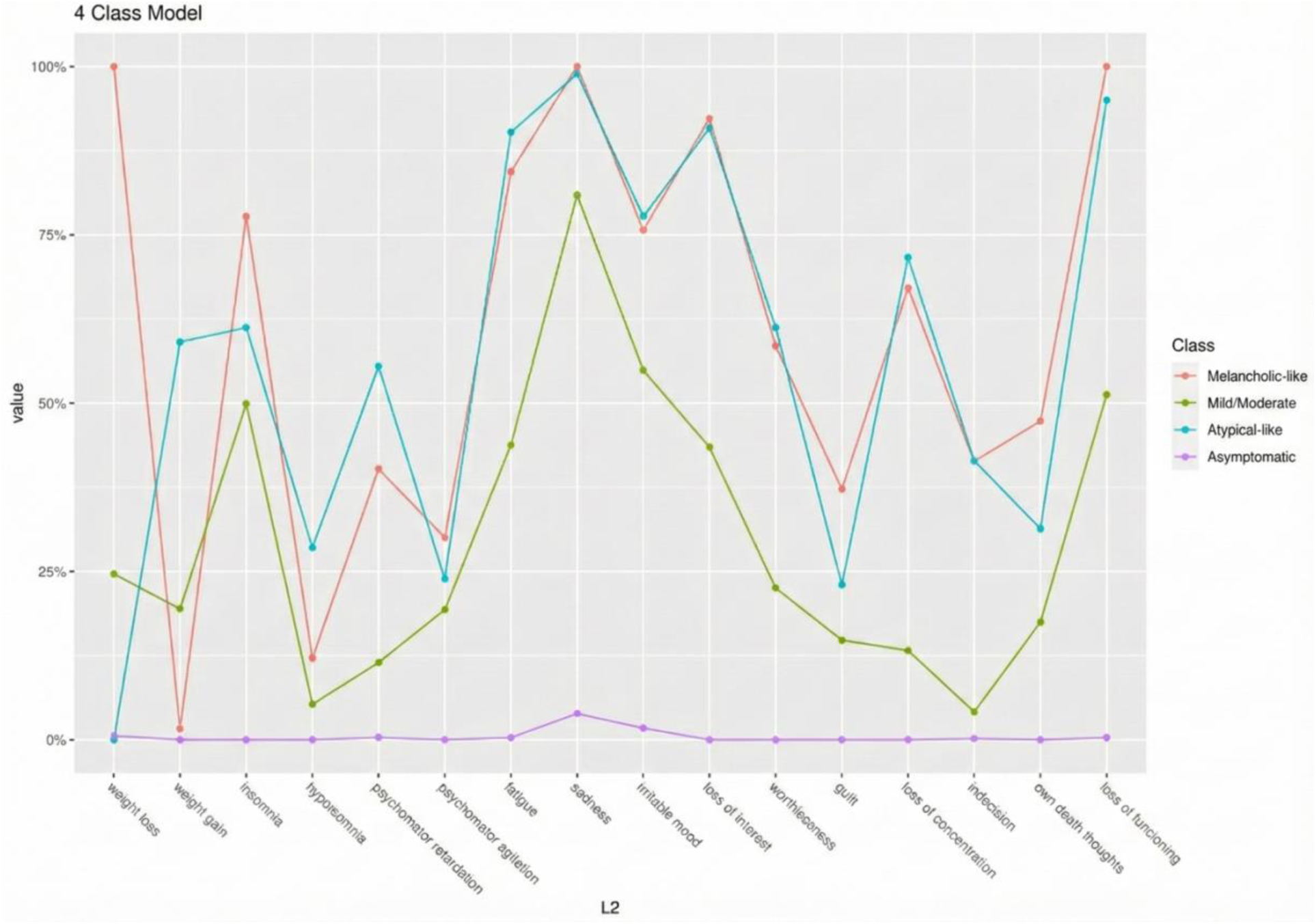
Symptom probability profiles for the four-class LCA models: asymptomatic, mild- moderate, atypical-like, and melancholic-like classes.

Applying DSM-IV criteria to SCID data, 78% of individuals in the atypical-like class and 88% in the melancholic-like class met criteria for a major depressive episode (MDE), compared with 17% of the mild–moderate class.

### Sociodemographic and immune-metabolic characteristics across classes

We next examined whether the latent classes differed in sociodemographic, clinical, and immune–metabolic characteristics (Table 2). Women were overrepresented across the sample and were particularly common in the two higher-severity classes. Current smoking was also more frequent in the atypical-like and melancholic-like classes than in the asymptomatic and mild–moderate classes. Age and education did not differ meaningfully across classes.

**Table 2.** Characteristics of participants (n=653)

|  | **Whole sample**<br>**(n=653)** | **Asymptomatic**<br>**(n=291)** | **Mild–Moderate**<br>**(n=125)** | **Atypical-like (n=109)** | **Melancholic-like**<br>**(n=128)** |
| --- | --- | --- | --- | --- | --- |
| CRP, mg/L<br>(median, range) | 1.50 (0.15–19.30) | 1.42 (0.15–19.30) | 1.98 (0.16–16.40) | 2.60 (0.16–18.10) | 1.095(0.15–14.70) |
| log-CRP<br>(mean, SD) | 0.50 (1.10) | 0.4752 (1.10) | 0.661 (1.09) | 0.804 (1.10) | 0.155 (1.00) |
| Age, years<br>(mean, SD) | 39.91 (10.60) | 39.86 (10.98) | 40.93 (10.31) | 37.98 (10.52) | 40.70 (9.93) |
| Education, years (mean,<br>SD) | 9.18 (4.05) | 9.175 (3.92) | 9.089 (3.89) | 9.477 (3.79) | 9.032 (4.69) |
| Sex, % male | 42.27% | 46.74% | 44.00% | 24.77% | 30.47% |
| Sex, % female | 57.73% | 53.26% | 56.00% | 75.23% | 69.53% |
| BMI, kg/m<sup>2</sup> (median, range) | 26.30 (16.24 –44.79) | 26.62 (16.85 –44.55) | 26.02 (16.40 –40.51) | 27.29 (18.34 – 44.79) | 24.89 (16.24 – 39.90) |
| BMI, kg/m<sup>2</sup><br>(mean, SD) | 26.89 (4.91) | 27.13 (4.92) | 27.02 (4.89) | 28.21 (5.33) | 25.09 (3.95) |
| Smoking, % non-smoker | 83.15% | 87.29% | 85.60% | 78.90% | 75.00% |
| Smoking, % current | 16.85% | 12.71% | 14.40% | 21.10% | 25.00% |
| HDL-C, mg/dL (mean, SD) | 47.43 (11.94) | 46.09 (11.14) | 47.44 (12.12) | 47.67 (12.64) | 50.18 (12.49) |
| LDL-C, mg/dL (mean, SD) | 122.20 (35.36) | 122.10 (34.03) | 122.47 (32.58) | 122.9 (36.46) | 121.62 (39.96) |
| Total cholesterol, mg/dL<br>(mean, SD) | 193.50 (46.44) | 193.3 (50.34) | 194.10 (39.95) | 194.20 (41.84) | 192.60 (47.29) |
| Fasting glucose, mg/dL<br>(mean, SD) | 89.10 (14.80) | 90.66 (17.36) | 90.18 (12.64) | 86.43 (12.66) | 86.29 (10.22) |
| Triglycerides, mg/dL<br>(median, IQR) | 95.00 (81.00) | 94.00 (82.50) | 99.00 (93.00) | 103.00 (74.00) | 84.5 (72.75) |

Median hs-CRP differed across the two higher-severity classes. hs-CRP was highest in the atypical-like class (median 2.60 mg/L) and lowest in the melancholic-like class (median 1.10 mg/L). Overweight or obesity, defined as BMI ≥25 kg/m², was more frequent in the atypical-like class (72.5%; 79/109) than in the melancholic-like class (47.7%; 61/128; χ²(1)=13.99, p<0.001).

### Biomarker associations with latent class membership

Multinomial logistic regression models were used to examine associations between immune– metabolic biomarkers and latent class membership, using the asymptomatic class as the reference group (Table 3). In Model 1, adjusted for age, sex, education, and smoking, the melancholic-like class was associated with lower log-CRP (OR 0.678, 95% CI 0.551–0.836). This association remained significant after additional adjustment for BMI in Model 2 (OR 0.768, 95% CI 0.612–0.965).

**Table 3.** Multinomial logistic models for biomarkers and latent classes.

| Biomarker | Model | Mild–moderate OR (95% CI) | Atypical-like OR (95% CI) | Melancholic-like OR (95% CI) |
| --- | --- | --- | --- | --- |
| CRP, log-transformed | Model 1 | 1.132 (0.926–1.385) | **1.255 (1.013–1.555)** | **0.678 (0.551–0.836)** |
|  | Model 2 | 1.163 (0.933–1.449) | 1.195 (0.943–1.513) | 0.768 (0.612–0.965) |
| HDL-C | Model 1 | 1.007 (0.988–1.026) | 1.000 (0.980–1.021) | 1.023 (1.004–1.042) |
|  | Model 2 | 1.008 (0.988–1.027) | 1.003 (0.983–1.024) | 1.017 (0.998–1.037) |
| LDL-C | Model 1 | 0.999 (0.993–1.006) | 1.003 (0.996–1.010) | 0.999 (0.992–1.005) |
|  | Model 2 | 0.999 (0.993–1.006) | 1.002 (0.995–1.009) | 1.000 (0.993–1.007) |
| Total cholesterol | Model 1 | 1.000 (0.995–1.005) | 1.002 (0.997–1.007) | 0.999 (0.994–1.004) |
|  | Model 2 | 1.000 (0.995–1.005) | 1.001 (0.996–1.007) | 1.000 (0.995–1.006) |
| Fasting glucose | Model 1 | 0.998 (0.982–1.014) | 0.987 (0.965–1.010) | 0.976 (0.955–0.997) |
|  | Model 2 | 0.999 (0.983–1.016) | 0.983 (0.960–1.007) | 0.986 (0.964–1.008) |
| Triglycerides, log-transformed | Model 1 | 0.984 (0.682–1.421) | 1.336 (0.904–1.975) | 0.704 (0.473–1.049) |
|  | Model 2 | 0.996 (0.670–1.480) | 1.168 (0.762–1.789) | 0.944 (0.613–1.452) |

The atypical-like class showed higher log-CRP in Model 1 (OR 1.255, 95% CI 1.013–1.555), but this association attenuated after BMI adjustment and was no longer statistically significant in Model 2 (OR 1.195, 95% CI 0.943–1.513). This suggests that the higher hs-CRP observed in the atypical-like class was at least partly accounted for by differences in BMI.

No robust associations were observed for LDL-C, total cholesterol, fasting glucose, or log-transformed triglycerides after covariate adjustment. HDL-C was positively associated with membership of the melancholic-like class in Model 1 (OR 1.023, 95% CI 1.004–1.042), but this association attenuated after BMI adjustment in Model 2 (OR 1.017, 95% CI 0.998–1.037).

## Discussion

In this population-based São Paulo cohort, latent modelling identified four depressive symptom profiles, including two high-severity phenotypes distinguished by opposing neurovegetative features. These clinical phenotypes were accompanied by divergent CRP profiles: atypical-like depression showed higher CRP that was largely accounted for by BMI, whereas melancholic-like depression showed lower CRP that persisted after BMI adjustment. These findings suggest that immune–metabolic differences in depression do not simply track diagnostic status or symptom severity but may cluster around specific symptom constellations even in a population exposed to substantial structural adversity.

Our approach directly addresses the heterogeneity inherent in the polythetic definition of MDD, which allows for numerous symptom combinations and may group biologically distinct conditions under a single diagnosis (3,4). By applying latent class analysis to DSM-IV symptoms, we move beyond this syndromic categorization. The resulting model not only identified a clear severity continuum but also isolated two distinct, high-severity classes whose symptom profiles - centred on weight gain or loss - naturally align with the classic atypical and melancholic syndromes. This demonstrates that data-driven methods can successfully parse the heterogeneity of the depressive syndrome into more homogeneous phenotypes, which our subsequent analysis found to be associated with divergent immune profile. Furthermore, this finding specifically converged with prior work from our team which showed similar differentiation (14).

Our findings also advance the field by demonstrating that inflammation is not uniformly elevated in depression but instead varies meaningfully across symptom-derived phenotypes. This is a critical refinement because much of the prior literature has treated MDD as a single, homogeneous entity when evaluating inflammatory biomarkers, contributing to inconsistent or weak associations. By contrast, our data show that distinct depressive subtypes carry distinct immunometabolic signatures, and that these signatures align with established clinical phenotypes: atypical-like depression is characterized by BMI-dependent inflammatory elevations, whereas melancholic-like depression presents with a BMI-independent low hs-CRP profile accompanied by relatively higher HDL-cholesterol levels. This pattern highlights that inflammation may not simply index overall depressive severity but instead tracks with specific neurovegetative constellations, particularly those related to appetite and weight change. Our findings agree with previous literature growing (62) support for the view that immune pathways contribute to only a subset of depressive presentations, particularly those involving reversed neurovegetative symptoms and increased adiposity.

The robustness of this association in our cohort strengthens this evidence. For the atypical-like subtype, our results present a more complex picture. While prominent studies associate this phenotype with elevated inflammation and metabolic dysregulation (13,29,50), our adjusted models showed that its inflammatory association was largely concurrent with higher BMI. This divergence does not refute the established literature but may reflect specific characteristics of our sample. Notably, our cohort had a high prevalence of overweight/obesity, which may have created a ceiling effect, attenuating the specific inflammatory signal of the atypical-like phenotype.

Identifying such subgroups is essential for precision psychiatry: anti-inflammatory or immunomodulatory interventions may hold therapeutic promise for individuals with inflammation-linked atypical-like depression but are unlikely to benefit - and may even be inappropriate for - individuals with melancholic-like profiles marked by physiological immune activity. Conversely, melancholic-like depression, with its BMI-independent low CRP levels, may reflect distinct biological pathways, such as heightened HPA-axis activation (61) or altered autonomic and metabolic dysregulation, suggesting the need for mechanistically different treatment strategies. Such biologically anchored phenotypes could also inform early identification strategies in primary care by highlighting symptom clusters that should prompt metabolic and inflammatory screening.

The findings have implications for biomarker stratification in depression but should be interpreted cautiously given the cross-sectional design and the limited biomarker panel. If replicated longitudinally, symptom-derived phenotypes may help identify subgroups in which immune-metabolic pathways are more relevant to disease expression or treatment response. BMI-related inflammatory elevation in atypical-like depression and BMI-independent lower hs- CRP in melancholic-like depression suggest that future intervention studies should not assume a uniform inflammatory mechanism across depressive presentations. Future work should examine longitudinal trajectories, treatment response, and mechanistic links e.g., HPA axis, microglial activation, and metabolic signalling across varying metabolic burdens.

### Limitations

Our study has several strengths, including the use of robust statistical methods, a relatively large sample thoroughly phenotyped by psychiatrists, and its status as one of the largest mental health studies with individuals from low- and middle-income countries where psychological stress is highly relevant. This is particularly relevant as most biomarker psychiatry studies are still dominated by high-income, lower-adversity, relatively selected samples. A limitation is that immune-metabolic disturbance was indexed by a relatively restricted biomarker panel. hs-CRP provides a useful marker of systemic low-grade inflammation, but it does not capture the full complexity of immune regulation, including cytokine signalling, cellular immune phenotypes, neuroendocrine pathways, or other stress-related disturbances. The cross-sectional design precludes causal inferences regarding associations between depressive subtypes and immune markers. Our sample consisted exclusively of residents of a large metropolitan area which may limit generalizability. We did not have information on the timing of the major depressive episode relative to biomarker assessment, which could affect the interpretation of CRP levels. Finally, our latent class analysis was based on DSM-IV symptom criteria. Although clinically relevant, this framework may not capture all symptom domains or other biopsychosocial factors relevant to heterogeneity.

## Conclusion

Latent depressive phenotypes showed divergent hs-CRP profiles in the São Paulo Megacity Mental Health Survey. Elevated hs-CRP in atypical-like depression was largely BMI-related, whereas melancholic-like depression showed a BMI-independent lower hs-CRP profile. These findings support the view that immune-metabolic biology in depression is phenotype-specific and may involve both higher and lower inflammatory profiles rather than a uniform increase in inflammation across depression.

## Author Contributions

IB, Y-PW, LAC and LHA designed the study. MCV and LHA wrote the protocol and created the database. BMC, DB-L, and GLS collected data. CHFA managed literature searches and summaries. CHFA and JMC-M performed statistical analyses under the supervision of LAC and LHA. CHFA, FCL, JMC-M, LAC, and LHA drafted the manuscript. All authors revised and approved the final manuscript.

## Data Availability

All data produced in the present study are available upon reasonable request to the authors

## Acknowledgments

The São Paulo Megacity Mental Health Survey is conducted in collaboration with the WHO World Mental Health Survey Initiative. We thank the WMH staff for assistance with instrumentation, fieldwork, and data analysis. The São Paulo Megacity Mental Health Survey (SPMHS) was funded by the Fundação de Amparo à Pesquisa do Estado de São Paulo (FAPESP; 03/00204-3) and the Conselho Nacional de Desenvolvimento Científico e Tecnológico (CNPq; 307623/2013-0). The SPMHS was carried out in conjunction with the World Health Organization World Mental Health (WMH). The main coordination centre activities, at Harvard University, were supported by the United States National Institutes of Mental Health (NIMH; R01-MH070884), the John D. and Catherine T. MacArthur Foundation, the Pfizer Foundation, and the U.S. Public Health Service (R13-MH066849, R01-MH069864, and R01- DA016558), as well as by the Fogarty International Center (FIRCA; R03-TW006481), the Pan American Health Organization (PAHO), the Eli Lilly and Company Foundation, Ortho-McNeil Pharmaceutical, GlaxoSmithKline, Bristol-Myers Squibb, and Shire. Funders had no role in study design, data collection/analysis, decision to publish, or manuscript preparation. None of the sponsors had any role in the design, analysis, interpretation of results, or preparation of this paper.

## Declaration of generative AI use in the manuscript preparation process

During the preparation of this work, the authors used DeepSeek to improve language clarity and readability, assist with consistency checks across the manuscript, and refine methodological descriptions and statistical reporting. After using this tool, the authors reviewed and edited the content as needed and take full responsibility for the content of the published article.

